# Low birthweight increased the risk of neonatal death twenty-folds in Northern Uganda: a community-based cohort study

**DOI:** 10.1101/2024.04.25.24306373

**Authors:** Beatrice Odongkara, Victoria Nankabirwa, Vincentina Achora, Anna Agnes Arach, Agnes Napyo, Milton Musaba, David Mukunya, Grace Ndeezi, Thorkild Tylleskär, James K Tumwine

## Abstract

**Background:** Low birthweight (LBW) is the leading cause of neonatal mortality and hospitalization worldwide. This study specifically aimed to: 1) determine the frequency of a) neonatal deaths and 2) assess their association with LBW in Northern Uganda.

**Methods:** A cohort study, nested in the Survival Pluss cluster randomized trial (NCT02605369), was conducted from January 2018 to February 2019 in Lira district, Northern Uganda. Out of 1877 pregnant women, 1556 live-born infants had their birthweight measured and were followed up to 28 days after birth. Generalized estimation equation regression models of the *Poisson* family with a log link were used to calculate the risk ratios between LBW and death.

**Results:** The risk of neonatal death was: 21/1,556 or 13.5 (95% CI: 8.8 – 20.6) per 1,000 live births. The respective sex and cluster adjusted proportion of neonatal death per 1000 live births among LBW, normal weight and not-weighed infants were 103 (95% CI: 47.2 – 212), 5.4 (95% CI: 2.1 – 13.9) and 167 (95% CI: 91.1-285). Compared to normal birthweight, LBW and not-weighed infants were each associated with a 20- and 30-folds increased risk of neonatal death.

**Conclusion:** In this community-based cohort study in Northern Uganda, neonatal mortality was 13.5/1000 live births. In the LBW and not-weighed groups, the risk of a neonatal death were more than twenty-times that of non-LBW infants. Efforts to reduce the number of LBW infants and/or prevent adverse outcomes in this patient group urgently are needed. In addition, all babies with should have birthweight recorded to facilitate early risk identification and management.

## Introduction

Of the 140 million infants born worldwide in 2014, an estimated 20 million (13%) were born with low birthweight (<2.5 kg).^1^ Ninety percent (18/20 million) of LBW infants were born in low- and middle-income countries (LMICs).^2^ In sub-Saharan Africa, LBW prevalence varied from 7.0% to 18.0%, with the highest prevalence observed in studies in areas with malaria in Tanzania.^3^ According to the Uganda Bureau of Statistics (UBOS) in 2011, some 10.4% of all live-born infants nationwide and 11.4% in the northern Uganda were LBW.^4^

LBW and preterm birth (PB) – a major component of LBW, are the main causes of neonatal and infant death, and are the second leading reason for under-five mortality globally.^5–7^ Nearly half (48%) of all neonatal deaths are due to complications of prematurity such as infections, respiratory distress syndrome, hypoglycaemia, or hypothermia.^8^

We hypothesised that LBW is associated with an increased risk of neonatal death compared to non-LBW. This study specifically aimed to: 1) determine the frequency of a) neonatal deaths and 2) assess their association with LBW in Northern Uganda.

## Subjects and methods

This was a cohort study nested within the Survival Pluss cluster randomized trial (ClinicalTrials.gov number NCT0260505369). The Survival Pluss study assessed the effect of an integrated package consisting of peer support by pregnancy buddies, provision of mama kits at household level (as opposed to health facility distribution) and mobile phone messaging on facility-based births. In the trial, pregnant women were enrolled at ≥ 28 weeks of gestation and followed through to delivery and 28 days postnatally (the neonatal period).

The study was conducted in Lira District, Northern Uganda from January 2018 to February 2019. Lira District had a population of about 400,000 people in 2010, dwelling in 13 sub-counties, one city and 751 villages. Lira district was chosen based on its being a post-conflict area with poor maternal and child health indicators, low proportion of health facility deliveries, high neonatal mortality, and limited data on LBW and PBs burden and associated risk factors.^9^ Three sub-counties were selected as study sites because they had the poorest maternal and child health indicators: Aromo, Agweng, and Ogur sub-counties.^9^ Each sub-county had at least one public health centre (HC), either level II (only outpatients), level III (having a maternity and in-patients) or level IV (having a surgical theatre). Each sub-county had one health centre with maternity (health centre, HC III or HC IV), and two additional lower-level health centres without maternity (HC II). Two of the HC IIIs (Agweng and Aromo), however, were not conducting deliveries before the project inception.

A total of 1877 mothers in 30 clusters were recruited into the trial at ≥28 weeks of gestation, followed up to birth and 28 days postnatally. We managed to obtain birthweight within two days after birth for 1,556 infants, Figure 1.

**Figure 1.**
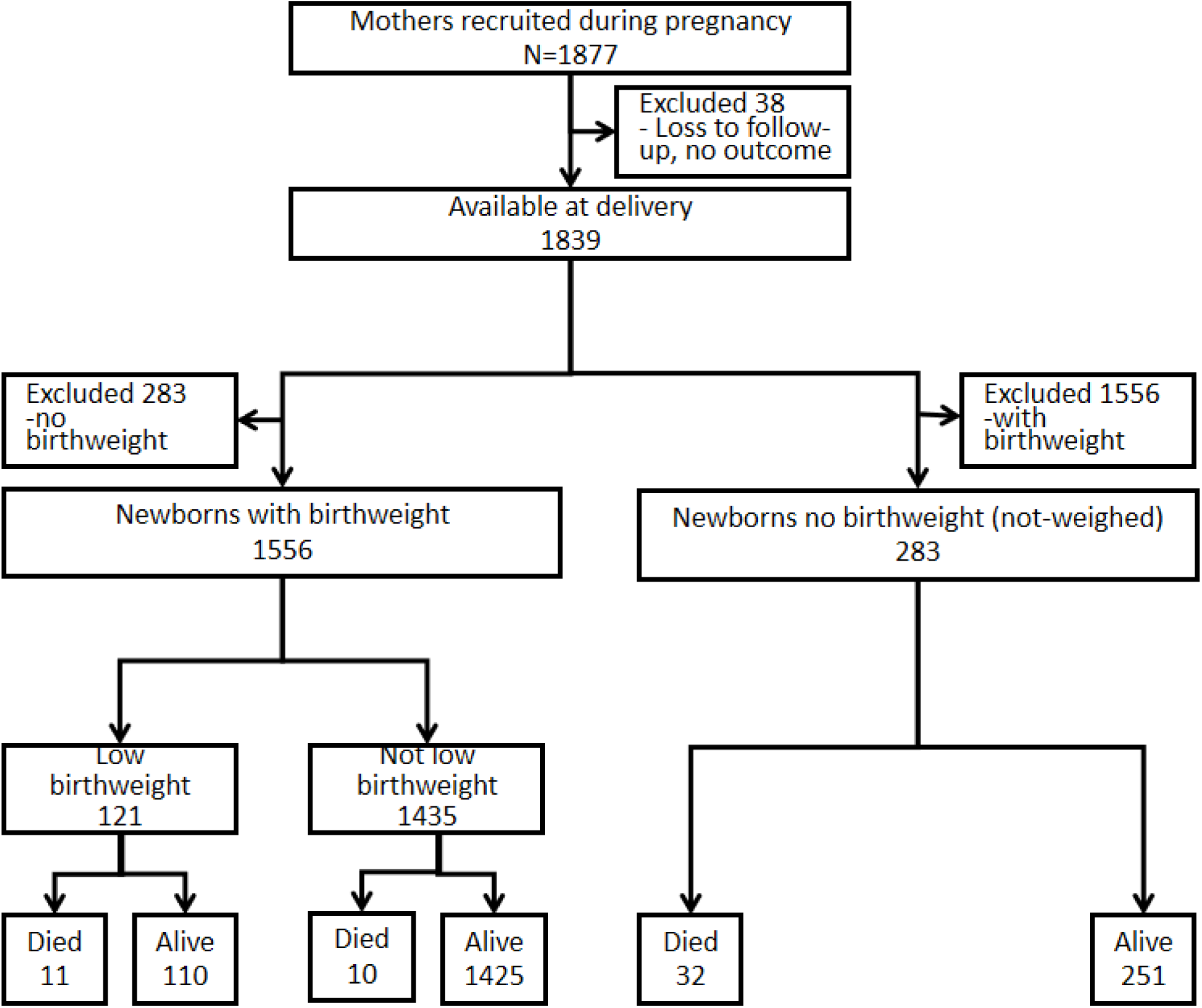
Study profile.

### Outcome

The primary outcome was neonatal death – defined as any demise of a live born infant within the first 28 days of life. Verbal autopsies were carried out for all deaths. The outcome was expressed per 1000 live births in the study cohort.

### Exposures

The primary exposure variable was low birthweight, while maternal demographic and clinical characteristics were covariates. A low birthweight (LBW) was defined as birthweight < 2.5kg measured within two days after birth.^10^ Birthweight was measured using a digital floor scale with mother/child function (seca, Hamburg, Germany), and recorded to the nearest 2 decimal points in kilograms. Education was recorded in years of completed schooling and dichotomized as 0–6 and 7, or more years in school. Wealth index quintiles were calculated based on key household assets and classified ranging from the 1 ‘poorest’ to 5 ‘wealthiest’ quintiles. This was further sub-grouped into three wealth groups: the lower 40% (1^st^ – 2^nd^ quintiles), the middle 40% (3^rd^ – 4^th^ quintiles), and the upper 20% (5^th^ quintile). Parity was the number of pregnancies the mother had before, and further re-categorised as ‘prime gravida (first time mother)’, ‘1–6’ and ‘7 or more’ children. Prior history of a small newborn was recorded as ‘yes’, if the mother had history of a small baby by her own assessment in her preceding pregnancy. The presence of maternal illnesses during pregnancy such as malaria or HIV were recorded as (‘yes’, ‘no’, or ‘unknown’), based on antenatal test results. Antenatal care (ANC) attendance was recorded as ‘yes’, if the woman attended antenatal clinic at least once during the current pregnancy. Intervention was recorded as ‘yes’, if the mother lived in an intervention cluster of the ‘parental trial’ and receiving the Survival Pluss intervention package (advice on birthplace by a peer buddy, SMS messages, and a clean birth kit ‘mama kit’) during pregnancy.

We analysed all 1839 of whom 1,556 mother-infant pairs from the Survival Pluss cohort of infants with birthweight. There was minimal difference in baseline characteristics between the analysed and excluded pairs, except for a few clinical factors, (Supplementary Table 1). The Survival Pluss study inclusion criteria were: mothers confirmed to be ≥28 weeks of gestation by last menstrual period or visibly pregnant, mothers who had no intention of moving away from the study area within a year of enrolment, and those without psychiatric illness, that could hinder the informed consent process. We excluded infants without birthweight which also included infants whose parents declined newborn examinations, infants that had died at birth or had severe congenital abnormalities (anencephaly and exomphalos).

### Study procedures

Prior to recruitment, research assistants were trained on the study protocol, weight measurement, electronic data collection tool, and the open data kit (ODK) software (https://opendatakit.org/). Pregnant mothers were identified by community recruiters, who informed the study team. The research assistants were then dispatched to see the identified mothers. Those who met the inclusion criteria were consented and recruited. The enrolled pregnant women were followed up to birth, and postnatally to 28 days. After birth, the same recruiters informed the study team, who in turn visited the mother-infant dyads at birth, for delivery questionnaire administration, and anthropometric measurements. The neonatal anthropometric (birthweight) was done within two days of postnatal life. The weighing scales were calibrated before each field visit, and before each measurement was taken. The weighing scales were checked for accuracy daily, with known standard weights. Data was collected using standardized pre-coded questionnaires in ODK, and immediately sent to the server for safe custody. Data cleaning and checking for completeness were done for quality control, throughout the data collection process. The principal investigator (BO) worked with and supervised the research assistants, on data collection and documentation.

### Statistical analysis

The data collected using ODK was sent to a server, from where it was downloaded to stata 14 (Stata Corp, College Station, Texas, US) for analysis. The proportions of neonatal death were presented as the number of each outcome measures (events), divided by the total number of live births reported per 1,000 live-born infants. To ensure that the reported death rate is not influenced unduly by the sex distribution or clustering effects in the data, making the results more generalizable and comparable, the neonatal death rates were adjusted for sex distribution and clustering effects, then expressed as the number of deaths per 1,000 live births, to provide a comparison across different study populations. Descriptive statistics for categorical variables were summarized into proportions, Table 1. The risk factors for neonatal death were analysed using bivariable and multivariable generalised estimation equation (GEE), for the binary categorical outcomes of death (Tables 2 respectively). Significant factors with p value ≤ 0.05 at bivariable analysis were taken into the multivariable GEE model, with a log link to Poisson family, adjusting for clustering and potential confounding. Known risk factors for neonatal death such as wealth index, and integrated intervention combinations, were also added into the final regression model even if they were not significantly associated with the adverse neonatal outcomes. The crude and adjusted risk ratios were compared during the multivariable regression analysis. A difference of ≥10% between crude and adjusted risk ratios were considered confounding.

**Table 1.** Baseline and clinical characteristics of participants in Lira district northern Uganda.

| Characteristics | All<br>N=1839 | Infants' Vital Status |  |
| --- | --- | --- | --- |
|  |  | Alive<br>n=1786 | Died<br>n=53 |
| Birthweight |  |  |  |
| Non-Low birthweight<sup>a</sup> | 1435(78.0) | 1425(79.8) | 10(18.9) |
| Low birthweight | 121(6.6) | 110(6.2) | 11(20.7) |
| Birthweight not measured | 283(15.4) | 251(14.0) | 32(60.4) |
| Maternal age groups in years |  |  |  |
| 12-19 | 504(27.4) | 473(26.5) | 31(58.5) |
| 20-34 | 1147(62.4) | 1131(63.3) | 16(30.2) |
| >=35 | 188(10.2) | 182(10.2) | 6(11.3) |
| Maternal education |  |  |  |
| 0-6 years | 1486(80.8) | 1442(80.7) | 44(83.0) |
| >=7 years | 353(19.2) | 344(19.3) | 9(17.0) |
| Marital status |  |  |  |
| Others | 166(9.0) | 161(9.0) | 5(9.4) |
| Married | 1673(91.0) | 1625(91.0) | 48(90.6) |
| Wealth index groups |  |  |  |
| Lower 40% | 824(44.8) | 801(44.8) | 23(43.4) |
| Middle 40% | 653(35.5) | 630(35.3) | 23(43.4) |
| Upper 20% | 362(19.7) | 355(19.9) | 7(13.2) |
| Paternal occupation |  |  |  |
| Unemployed | 167(9.2) | 165(9.4) | 2(4.7) |
| Employed | 388(21.5) | 380(21.5) | 8(18.6) |
| Farmer | 1251(69.3) | 1218(69.1) | 33(76.7) |
| Domestic water source |  |  |  |
| Other | 674(36.7) | 649(36.3) | 25(47.2) |
| Tap/Borehole | 1165(63.3) | 1137(63.7) | 28(52.8) |
| Intervention |  |  |  |
| No | 872(47.4) | 847(47.4) | 25(47.2) |
| Yes | 967(52.6) | 939(52.6) | 28(52.8) |
| History of a small newborn |  |  |  |
| Prime gravida | 420(22.8) | 395(22.1) | 25(47.2) |
| Yes | 294(16.0) | 282(15.8) | 12(22.6) |
| No | 1125(61.2) | 1109(62.1) | 16(30.2) |
| Parity<sup>*</sup> |  |  |  |
| Prime Gravida | 420(22.8) | 395(22.1) | 25(47.2) |
| 1-6 | 1232(67.0) | 1207(67.6) | 25(47.2) |
| 7 or more | 187(10.2) | 184(10.3) | 3(5.7) |
| Maternal HIV |  |  |  |
| Negative | 1675(91.1) | 1629(91.2) | 46(86.8) |
| Positive | 83(4.5) | 81(4.5) | 2(3.8) |
| Unknown | 81(4.4) | 76(4.3) | 5(9.4) |
| Antenatal attendance |  |  |  |
| No | 386(21.0) | 374(20.9) | 12(22.6) |
| Yes | 1453(79.0) | 1412(79.1) | 41(77.4) |
| IPT for Malaria in pregnancy |  |  |  |
| No | 761(41.4) | 751(42.1) | 10(18.9) |
| Yes | 1078(58.6) | 1035(57.9) | 43(81.1) |
| Maternal malaria in pregnancy |  |  |  |
| No malaria | 581(31.6) | 559(31.3) | 22(41.5) |
| Yes | 453(24.6) | 445(24.9) | 8(15.1) |
| Unknown | 805(43.8) | 782(43.8) | 23(43.4) |
| Home delivery |  |  |  |
| No, Facility Births | 1209(65.7) | 1173(65.7) | 36(67.9) |
| Yes, Home Births | 630(34.3) | 613(34.3) | 17(32.1) |
| Infant sex |  |  |  |
| Female | 878(48.9) | 855(49.0) | 23(43.4) |
| Male | 919(51.1) | 889(51.0) | 30(56.6) |

*IPT* intermittent preventive treatment for malaria, *HIV* Human immunodeficiency virus, *N/n (%)* frequency with percentages in parentheses. <sup>a</sup> either because of lack of consent to assess the child, or because the study team did not reach the family within 48 hours after birth or the child had a major malformation or had already died before the arrival of the study team

### Ethical considerations

Ethical clearance was obtained from Makerere University School of Medicine Research and Ethics Committee (SOMREC no. 2015/085), the Uganda National Council for Science and Technology (UNCST no. HS 2478) and REK Vest in Norway (No. 2018/58/REK Vest). Permission was obtained from the district and health facility administrations. The study was also registered with ClinicalTrial.gov NCT02605369). Written informed consent was obtained from each Survival Pluss study participant. Participant confidentiality was maintained, through use of password protected mobile phones and computers.

## Results

### Study profile

A total of 1,877 pregnant women were recruited into the Survival Pluss trial. Forty-four were lost to follow-up before birth and some were not weighed within two days of birth. Of the 1,556 mother-infant dyads with birthweight, 121 or 7.8% had low birthweight, 21 or 1.3% died and 32 or 2.1% were hospitalized during the neonatal period. Only 2/21 or 9.5% of infants who died were actually hospitalized, (Figure 1) of whom one was a home birth. Noteworthy, 14/21 or 67% of neonatal deaths occurred among infants born in health facilities and 7 (33%) among home births. Of the 21 neonatal deaths, 11 (>50%) were low birthweight. Of these 10 (91%) were non-hospitalised and died at home including the 7/11 born in health facilities.

### Baseline demographic and clinical characteristics of study participants

A total of 1839 of the 1877 infants were followed up to delivery. We obtained birthweight for 1556 of 1877 or 85% mother-infant dyads, 85% of the total. Of the 1839 mother-infant dyads, a quarter (504 OR 27.4%) of the mothers were first time mothers (prime gravida); 1.4% were twins, and 91% were married, Table 1. Most of the fathers (69%) were subsistence farmers. Most families (63%) used tap or borehole water for domestic consumption. About 4.5% of the mothers were HIV infected, while up to 4.4% did not know their HIV status. Close to 14% of mothers had history of a small new-born in the most recent (second last) delivery. The male to female ratio was close to 1:1, Table 1.

### Neonatal deaths

Of the 89 deaths 38 were stillborn, half of the neonatal deaths occurred on the first day of life, Figure 2. The neonatal deaths decreased with increasing postnatal age.

**Figure 2.**
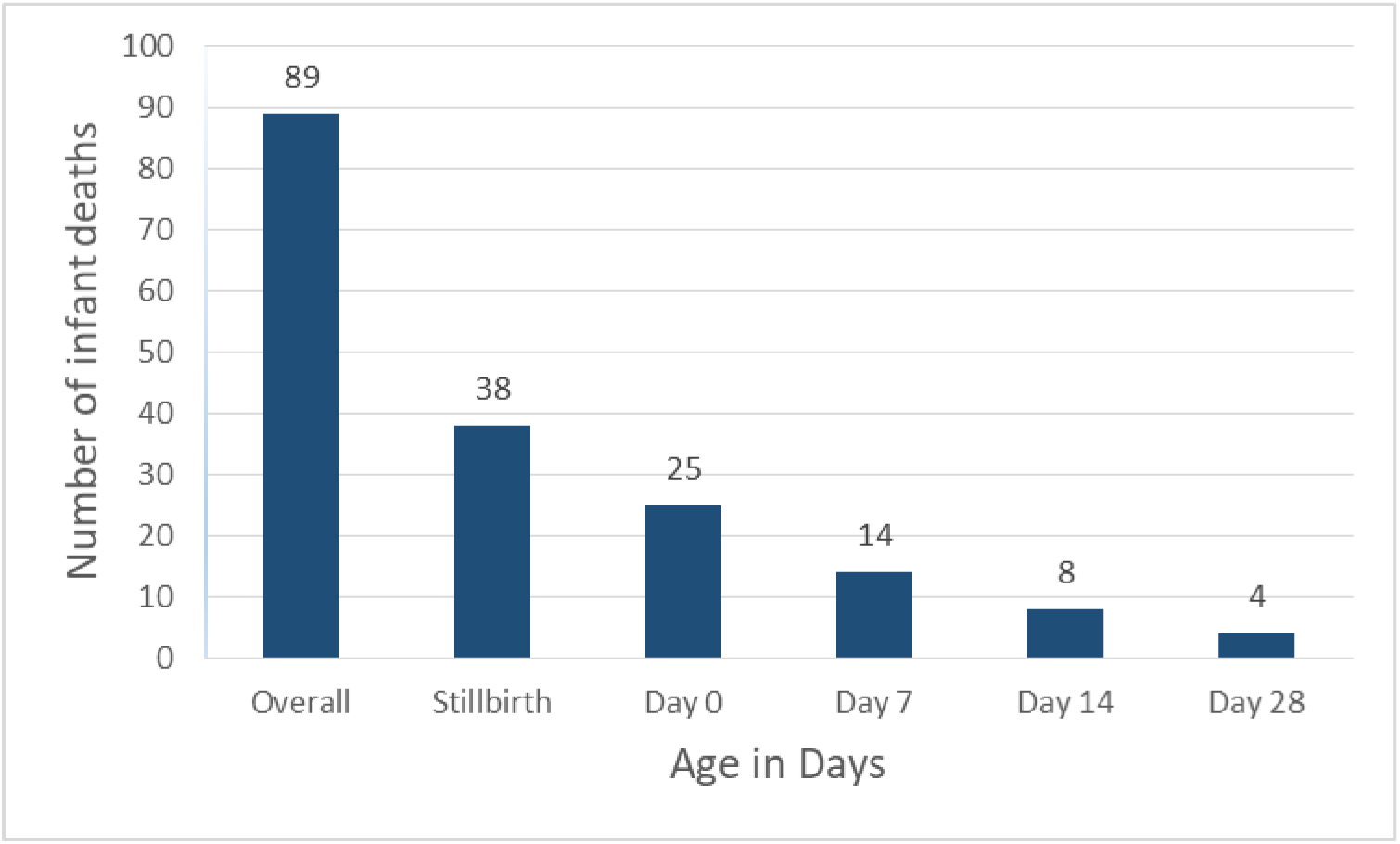
Number of neonatal deaths.

### The proportion of neonatal deaths

The number with respective crude and adjusted risks (proportions) of neonatal deaths (mortality) are summarized in Table 2. The overall proportion of neonatal death in this cohort was 32.6 (96% CI: 20.9 – 50.6) per 1,000 live births. The observed proportions of neonatal death among the low birthweight was 103 (95% CI: 47.2 – 212) and among non-low birthweight was 5.4 (95% CI: 2.1 – 13.9) per 1,000 live births. An estimated 167 un-weighed infants died per 1,000 live birth in the neonatal period, Table 3. Compared to the non-low birthweight, the un-weighed infants had 30-while the low birthweight had 20-times higher mortality rates.

**Table 2.**
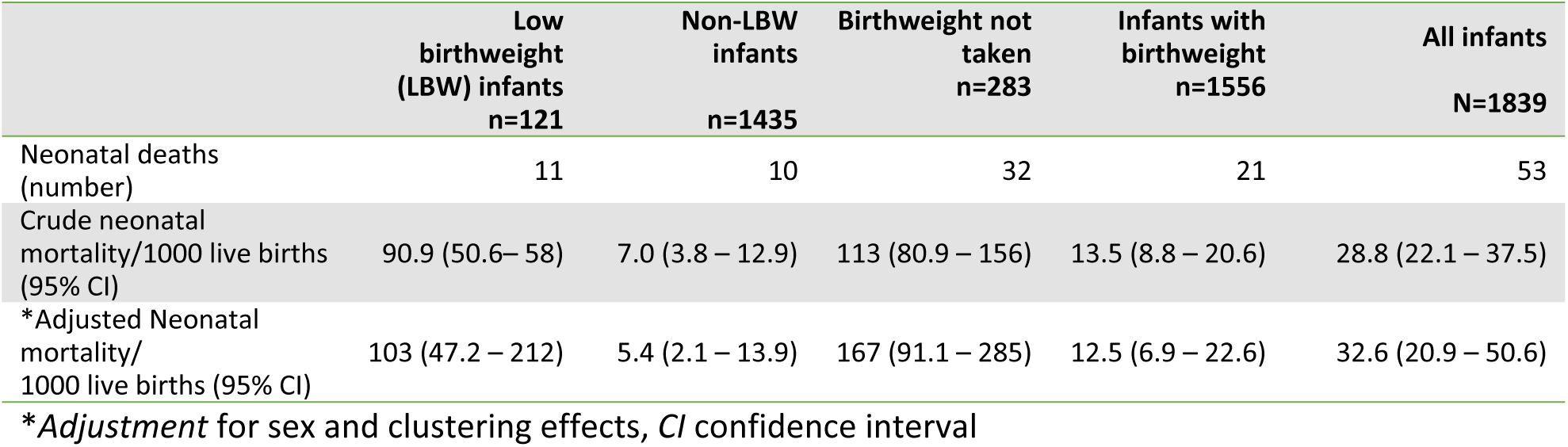
Numbers and adjusted rates of neonatal deaths.

|  | Low birthweight (LBW) infants<br>n=121 | Non-LBW infants<br>n=1435 | Birthweight not taken<br>n=283 | Infants with birthweight<br>n=1556 | All infants<br>N=1839 |
| --- | --- | --- | --- | --- | --- |
| Neonatal deaths (number) | 11 | 10 | 32 | 21 | 53 |
| Crude neonatal mortality/1000 live births (95% CI) | 90.9 (50.6– 58) | 7.0 (3.8 – 12.9) | 113 (80.9 – 156) | 13.5 (8.8 – 20.6) | 28.8 (22.1 – 37.5) |
| *Adjusted Neonatal mortality/1000 live births (95% CI) | 103 (47.2 – 212) | 5.4 (2.1 – 13.9) | 167 (91.1 – 285) | 12.5 (6.9 – 22.6) | 32.6 (20.9 – 50.6) |

\*Adjustment for sex and clustering effects, CI confidence interval

**Table 3.** Bi- and multi-variable analysis for risk factors for neonatal deaths in Lira district.

**Tab 3. Bi- and multi-variable analysis for risk factors for neonatal deaths in Lira district**
| Characteristics | All<br>N=1839 | Vital<br>status<br>Died<br>n=53 | Crude RR<br>(95% CI)<br>N=1839 | p<br>value | Adjusted RR<br>(95% CI)<br>N=1839 | p<br>value |
| --- | --- | --- | --- | --- | --- | --- |
| *Birthweight* |  |  |  |  |  |  |
| Non-Low birthweight | 1435(78.0) | 10(18.9) | Ref |  |  |  |
| Low birthweight | 121(6.6) | 11(20.7) | 13.2(6.6 – 26.1) | 0.000 | 13.2(6.9 – 25.3) | 0.000 |
| Birthweight not measured | 283(15.4) | 32(60.4) | 17.0(9.1 – 32.0) | 0.000 | 15.8(8.4 – 29.8) | 0.000 |
| *Maternal age groups in years<sup>‡</sup>* |  |  |  |  |  |  |
| 12-19 | 504(27.4) | 31(58.5) | 4.3(2.3 – 8.0) | 0.000 | 3.1(1.3 – 7.4) | 0.012 |
| 20-34 | 1147(62.4) | 16(30.2) | Ref |  |  |  |
| >=35 | 188(10.2) | 6(11.3) | 2.2(0.9 – 5.3) | 0.088 | 2.3(0.5 – 10.1) | 0.253 |
| *Maternal education* |  |  |  |  |  |  |
| 0-6 years | 1486(80.8) | 44(83.0) | Ref |  |  |  |
| >=7 years | 353(19.2) | 9(17.0) | 0.8(0.5 – 1.5) | 0.544 |  |  |
| *Marital status* |  |  |  |  |  |  |
| Others | 166(9.0) | 5(9.4) | Ref |  |  |  |
| Married | 1673(91.0) | 48(90.6) | 1.0(0.4 – 2.3) | 0.947 |  |  |
| *Wealth index groups* |  |  |  |  |  |  |
| Lower 40% | 824(44.8) | 23(43.4) | Ref |  |  |  |
| Middle 40% | 653(35.5) | 23(43.4) | 1.3(0.8 – 2.0) | 0.268 | 1.1(0.7 – 1.7) | 0.636 |
| Upper 20% | 362(19.7) | 7(13.2) | 0.7(0.3 – 1.6) | 0.378 | 0.8(0.3 – 1.7) | 0.498 |
| *Paternal occupation* |  |  |  |  |  |  |
| Unemployed | 167(9.2) | 2(4.7) | Ref |  |  |  |
| Employed | 388(21.5) | 8(18.6) | 1.8(0.6 – 6.9) | 0.417 |  |  |
| Farmer | 1251(69.3) | 33(76.7) | 2.3(0.7 – 7.4) | 0.177 |  |  |
| *Domestic water source* |  |  |  |  |  |  |
| Other | 674(36.7) | 25(47.2) | Ref |  |  |  |
| Tap/Borehole | 1165(63.3) | 28(52.8) | 0.6(0.4 – 1.0) | 0.073 | 0.7(0.4 – 1.1) | 0.046 |
| *Intervention* |  |  |  |  |  |  |
| No | 872(47.4) | 25(47.2) | Ref |  |  |  |
| Yes | 967(52.6) | 28(52.8) | 1.0(0.5 – 1.9) | 0.988 |  |  |
| *History of a small newborn<sup>‡</sup>* |  |  |  |  |  |  |
| Prime gravida | 420(22.8) | 25(47.2) | 1.5(0.8 – 2.8) | 0.220 | 1.8(0.9 – 3.7) | 0.116 |
| Yes | 294(16.0) | 12(22.6) | Ref |  |  |  |
| No | 1125(61.2) | 16(30.2) | 0.4(0.1 – 0.9) | 0.020 | 0.6(0.2 – 1.5) | 0.246 |
| *Parity<sup>‡</sup>* |  |  |  |  |  |  |
| Prime Gravida | 420(22.8) | 25(47.2) | 3.0(1.7 – 5.2) | 0.000 | 2.8(1.7 – 4.8) | 0.000 |
| 1-6 | 1232(67.0) | 25(47.2) | Ref |  |  |  |
| 7 or more | 187(10.2) | 3(5.7) | 0.8(0.3 – 2.7) | 0.751 | 0.8(0.3 – 2.8) | 0.777 |
| *Maternal HIV infection* |  |  |  |  |  |  |
| Negative | 1675(91.1) | 46(86.8) | Ref |  |  |  |
| Positive | 83(4.5) | 2(3.8) | 0.9(0.2 – 3.3) | 0.883 | 1.4(0.3 – 5.5) | 0.661 |
| Unknown | 81(4.4) | 5(9.4) | 2.3(0.8 – 6.8) | 0.104 | 1.1(0.2 – 2.1) | 0.915 |
| *Antenatal attendance* |  |  |  |  |  |  |
| No | 386(21.0) | 12(22.6) | Ref |  |  |  |
| Yes | 1453(79.0) | 41(77.4) | 0.9(0.5 – 1.7) | 0.729 |  |  |
| *Maternal malaria in pregnancy* |  |  |  |  |  |  |
| No malaria | 581(31.6) | 22(41.5) | Ref |  |  |  |
| Yes | 453(24.6) | 8(15.1) | 0.4(0.2 – 1.2) | 0.101 | 0.4(0.2 – 1.1) | 0.064 |
| Unknown | 805(43.8) | 23(43.4) | 0.8(0.4 – 1.3) | 0.298 | 0.5(0.3 – 0.9) | 0.022 |
| *Home delivery* |  |  |  |  |  |  |
| No, Facility Births | 1209(65.7) | 36(67.9) | Ref |  |  |  |
| Yes, Home Births | 630(34.3) | 17(32.1) | 1.0(0.5 – 1.7) | 0.877 |  |  |
| *Infant sex* |  |  |  |  |  |  |
| Female | 878(48.9) | 23(43.4) |  |  |  |  |
| Male | 919(51.1) | 30(56.6) | 1.2(0.7 – 2.2) | 0.451 |  |  |

<sup>‡</sup> A separate multivariable model with malaria in pregnancy instead of IPT in was done. <sup>\*</sup> Similarly, a separate model was also run for parity without history of a small newborn. IPT intermittent preventive treatment for malaria, HIV Human immunodeficiency virus, N/n(%) frequency with percentages in parentheses

### Risk factors for neonatal death in post-conflict Northern Uganda

Compared to normal birthweight (non-LBW) infants, the risk of neonatal death was significantly higher among LBW infants, Table 3. LBW infants were almost 13.2 times at more risk of dying in the neonatal period compared to non-LBW infants while the non-weighted infants were 15.8 times at risk of neonatal deaths than the non-low birthweight counterparts. Other factors that were associated with high likelihood of neonatal death include: maternal age 12 – 19 years (teenage motherhood), and unknown malaria status in pregnancy. Infants born to teens and first time mothers were each at thrice the risk of neonatal deaths than mothers aged 20-34 year and mothers with 1-6 children, Table 3.

## Discussion

The study from post-conflict northern Uganda provides important insights into neonatal mortality rates and the associated risk factors. In our cohort, the overall neonatal mortality risk was 28.8 per 1,000 live births while among infants with birthweight it was 12.5 per 1,000 live births. Lastly, the respective neonatal mortality among low-, non-LBW and un-weighed infants were 103, 5.4 and 167 per 1,000 live births. Compared to the normal birthweight, low birthweight and un-weighed infants were 20- and 30-times at higher risk of neonatal deaths. The findings indicate a significantly higher mortality rate among low birthweight (LBW) and un-weighed infants compared to non-LBW infants. Teenage motherhood and prime-parity were associated with an increased, while unknown maternal malaria status in pregnancy was associated with a reduced risk of neonatal deaths. Whereas we could not explain the reduced risk of neonatal death among mothers with unknown malaria status compared to those without malaria in pregnancy we discuss the rest of the findings in details hereunder.

The observed, overall low proportion of neonatal deaths in the cohort of infants with birthweight could be due to the attention given by the trial staff in the main trial. For instance, when staff identified sick infants during the new-born examination, they managed (PI) and/or referred them to health facilities for further care. Infants found to be having a low blood glucose were immediately encouraged to breastfeed and a repeat blood glucose was done to verify correction of the hypoglycaemia. Noteworthy, ≥90% of the neonatal deaths occurred outside health facilities, irrespective of place of birth. The low neonatal deaths rates in non-low birthweight infants confirms the protective effect of normal birthweight on neonatal deaths.

The overall population based neonatal deaths rates, and deaths among low birthweight and un-weighed infants were much higher than the sustainable development goal 3.2 target of <10 per 1,000 live births by 2030.^11^ The high neonatal mortality rates is similar to the national and low resource settings rates.

But, LBW infants were 20-times and un-weighed infants 30-folds more likely to die compared to non-LBW infants. In fact, half of the neonatal deaths occurred in the LBW group which constitutes less than 8% of the whole population of babies with birthweight. This means that LBW is one of the main drivers for the neonatal mortality. If this easily identifiable risk group for neonatal deaths could receive more attention from the health systems, we could potentially reduce neonatal mortality drastically. The finding that LBW is associated with an increased risk of neonatal death is not unique to our setting, with additional documented evidence across the globe.^12^

The possible explanations for the increased risk of neonatal death with LBW may include: the well-known associations of LBW with hypothermia, birth asphyxia, and hypoglycaemia.^13^ Another explanation for the link between LBW and neonatal adverse outcomes in our cohort may be that more than 50% of neonatal deaths occurred among non-hospitalised LBW infants. The majority of these were born in health facilities and discharged home immediate post-delivery. Implications – all low birthweight infants irrespective of birth place needs to receive the attention of the health system, for instance in the form of assistance with kangaroo mother care (KMC), and breastfeeding.

Moreover, infants without birthweight had more than 30-folds risks of neonatal death compared to non-low birthweight infants. The reasons for these could be that un-weighed babies who died before the PI (BO) and her research team of health workers could reach and assess them for severe illnesses needing further care.

Furthermore, we also report increased risk of neonatal deaths among teenage and first time mothers compared to those aged 20-34 years and 1-6 pregnancies respectively. Teenage pregnancy has long been known to be associated with adverse pregnancy outcomes such as preterm births, low birthweight and neonatal deaths worldwide.^14^ In addition, in this dataset, teenage mothers were also the majority first time mothers.

Without interventions to accelerate the decline in infant mortality, especially among LBW infants, an estimated 31 million infants will die in the early infancy, by the year 2030.^15,16^ A number of cost-effective interventions exist to identify (weigh all the infants at birth) and reduce LBW and its component PBs.^17^ This may then reduce complications, and improve survival, in order to reach the 2030 Sustainable Development Goal number 3 (SDG 3) target of below 12 neonatal deaths per 1,000 live births in each country.

## Limitations and strengths

Our inability to assess LBW related neonatal deaths among infants with missing birthweight and those who delivered before inclusion in the trial is a limitation to this study. It is possible that these mothers had worse outcomes than what we have reported.

This study however, had several strengths. Firstly, it was a community-based cohort – likely to reflect the community at large. Secondly, we were able to follow-up and obtain birthweight within 48 hours on 85% of the cohort, minimising the risk of selection bias. Lastly, the mothers were interviewed shortly after the events, thereby minimising the risk of recall bias.

## Conclusion

In this community-based cohort study in Northern Uganda, the neonatal mortality was 12.5/1000 live births. Half of the neonatal deaths among infants with birthweight occurred in the LBW group which constitutes less than 8% of the whole population and in this group, the risk of a neonatal death was almost 20-fold compared to non-LBW infants. Un-weighed infants even had a higher risk of dying compared to the non-low birthweight counterparts. Infants of teenage and first time mothers had more than twice the risk of neonatal deaths compared to mothers aged 20-34 years.

## Recommendation

Given the high risk of morbidity and mortality among LBW and un-weighed infants, we recommend scaling up of available proven interventions such as taking birthweight for all babies irrespective of place of birth and kangaroo mother care, etc., along the continuum of care to prevent LBW and preterm birth and their related complications in this community.

## Data Availability

Unrestricted data access can be obtained from the corresponding author on reasonable request.

## Declarations

Ethics approval and consent to participate: approvals were obtained from UNCST. Informed consent and ascent obtained from study participants. Confidentiality was ensured through use of identification numbers, password protected computers and storage in lockable office spaces.

Consent for publication: All authors consented to the publication submission Availability of data and materials: Data will be availed on reasonable requests Competing interests: We declare no competing interests

Funding: NORHED funded the study in totality

Authors’ contributions: All authors contributed to the study Acknowledgements: Study participants and research assistants

